# The dimensionality of the Death Anxiety Inventory-Revised in Colombian-aged adults

**DOI:** 10.1101/2024.02.27.24303479

**Authors:** Adalberto Campo-Arias, Monica Reyes-Rojas, Guillermo Augusto Ceballos-Ospino

## Abstract

The Death Anxiety Inventory-Revised (DAI-R) is a relatively new instrument to quantify anxiety about death in different contexts. However, the dimensionality in the elderly population is unknown. The study aimed to corroborate the dimensionality of the DAI-R among Colombian-aged adults. A psychometric study was conducted with the participation of 100 aged adults (M=68.82, SD=4.82; 52% were male gender). This scale has 17 items, grouped into four dimensions or factors, and a dichotomous answer pattern. Dimensionality was tested using confirmatory factor analysis (CFA), and goodness-of-fit indicators were computed. The CFA showed unacceptable goodness-of-fit indicators, and the four-dimensional structure of the DAI-R was rejected. In conclusion, the DAI-R has an unsatisfactory four-dimensional structure among Colombian-aged adults. Further research should corroborate this finding with a large sample size.

---

Death anxiety can be a natural or normative expectation about awareness of death, dying, or non-existence (Barrett, 2013; Lehto & Stein, 2009). However, death anxiety may become distressing if it involves excessive avoidance of situations associated with disease and death, disabling worry, and decreased life enjoyment (Furer & Walker, 2008). Then, distressing death anxiety refers to negative or maladaptive emotions, cognitions, or behaviors related to awareness of death, dying, or non-existence (Barrett, 2013; Lehto & Stein, 2009).

Death anxiety has been studied and measured from different theoretical models, such as terror management, existential, and integrated models (Furer & Walker, 2008; Iverach et al., 2014; Lehto & Stein, 2009). Recently, Zuccala et al. (2022) systematically reviewed the psychometric properties of 21 self-report death anxiety scales and six subscales and reported that most of them have strong evidence of validity and reliability. However, among Spanish speakers, one of the most used scales to measure death anxiety is the Death Anxiety Inventory-Revised, DAI-R (Tomás-Sábado et al., 2005). The DAI-R is based on the idea that the construct consists of external and internal factors in a multidimensional model by Templer (1970, 1971). Internal factors include age, gender or religious beliefs, and physical and mental health. External factors include the presence of medical problems, stressful environments, life-threatening experiences, history of loss of loved ones, and other sociocultural aspects (Adelirad et al., 2021; Alvarado et al., 1995; Lehto & Stein, 2009; Missler et al., 2012; Templer, 1970; 1971). Nevertheless, Templer’s scale’s 7-point Likert-type answer pattern could be problematic for its Spanish version, in which answer options between two and five usually work well (Lopez, 2005).

The Death Anxiety Inventory (DAI) was developed in Spanish (Tomás-Sábado et al., 2005); an English version was later available (Tomás-Sábado et al., 2005). The DAI is a 20-item self-administered instrument that can be administered with Likert scales (six options) or dichotomous items (true or false) distributed across five dimensions: “Externally generated death anxiety,” “meaning and acceptance of death,” “thoughts about death,” “life and death,” and “shortness of life” (Tomás-Sábado & Gómez-Benito, 2005).

However, later studies showed that the DAI had several items that showed a poor correlation with the total score and inconsistency with the factor loadings of the five dimensions (Limonero et al., 2003; Tomás-Sábado & Limonero, 2004). Therefore, the 17-item version (DAI-R) was introduced in which items associated with life span and aging were omitted. The DAI-R is composed of four factors, namely, “acceptance of death,” “externally generated death anxiety,” “finality of death,” and “thoughts about death” (Tomás-Sábado et al., 2005).

The “acceptance of death” dimension refers to personal understanding, the experience of fear or not towards this universal event, and taking responsibility for life. The dimension “externally generated death anxiety” is related to aspects outside the person’s voluntary control, such as illness, knowledge of death, and cultural elements. The “finality of death” dimension refers to the conception of death as a positive or negative event that grants or restricts the meaning of life and is associated with the intention of life because human beings seek to give purpose to their existence and alleviate the fear of death. The “thoughts about death” dimension makes up the cognitive component of the construct by exploring attitudes, awareness of the possibility of one’s own or others’ death, and beliefs about the proximity of death or a long existence (Tomás-Sábado et al., 2005).

Information on the dimensionality of the DAI-R is scarce. Waite et al. (2022), among 2,205 adults from the general community in the United Kingdom between 18 and 83 years of age, observed that the five-option Likert version of the DAI-R presented an adequate structure in four dimensions, with a Cronbach’s alpha of .94 for the full DAI-R; however, they included among the participants a high proportion of people under 60 years of age. Also, the authors omitted to report Cronbach’s alpha for each dimension of the DAI-R. It is inappropriate to report global Cronbach’s alpha for multidimensional scales (Tavakol & Dennick, 2011).

The dimensional structure of health measurement scales must be corroborated in different populations, given that the dimensionality of psychometric instruments can be affected by the characteristics of the participating people (Keszei et al., 2010). In addition, a continuous process of review and refinement of health scales is necessary to guarantee the validity and reliability of the measurement (Campo-Arias & Pineda-Roa, 2022).

In the current study, the dimensionality of the DAI-R is described in a sample of older adults, and the internal consistency is reported separately for each of the four dimensions proposed for the DAI-R. The internal consistency of each dimension, previously forgotten in previous studies, is reported (Tomás-Sábado et al., 2005; Waite et al., 2022). The internal consistency of each dimension must be calculated for a multidimensional scale (Tavakol & Dennick, 2011). Besides, a Latin American sample with a differential cultural background is analyzed, such as the high frequency of older adults who live with other family members and have more outstanding community support and social participation (Esteve et al., 2022).

With older adulthood, people approach this natural and universal event of dying. Consequently, the perception of death usually changes due to the increasing and usual morbidity or deterioration in the quality of life that makes death more likely or imminent. Consequently, this can modify the instrument’s overall response pattern and dimensionality (Duran-Badillo et al., 2020). Most nurses are uncomfortable with death and dying (Gillan et al., 2014). However, these personnel, particularly in geriatrics, must be aware of the attitude towards death in service users. Holistic care requires the guidance of patients in coping with anxieties or fears regarding death (Eliopoulos, 2018).

The study’s objective was to corroborate the dimensionality of the DAI-R in older adults residing in Santa Marta, Colombia.

## Methods

### Design

A psychometric study was designed to evaluate the dimensionality of a health measurement instrument.

### Participants

A convenience sample of 100 older adults between 62 and 86 years old (68.82±4.82) was taken. The highest percentage were men, in a stable relationship (married or in a common-law union), with paid employment or receiving a pension, residing in low-income neighborhoods, and with a secondary education or less—more details of demographic characteristics in Table 1.

**Table 1.** Demographic characteristics of the participants.

| **Variable** | **Frequency and %** |
| --- | --- |
| Gender |  |
| *Female* | 48 |
| *Male* | 52 |
| Marital status |  |
| *Married or common-law union* | 60 |
| *Single, widowhood, separation, or divorce* | 40 |
| Paid employment or receiving a pension |  |
| *Yes* | 45 |
| *No* | 55 |
| Education |  |
| *High-school or less* | 65 |
| *College or university* | 35 |
| Income |  |
| *Low* | 79 |
| *High* | 21 |

### DAI-R

The DAI-R is made up of 17 items, as mentioned above, distributed in four dimensions or factors: “Death acceptance” (items 2, 3, 5, 6, 7, and 15), “externally generated death anxiety” (items 1, 8, 12, and 16), “death finality” (items 4, 9, 10 and 13) and “thoughts about death” (items 11, 14, and 17). The scores have a straightforward interpretation: the higher the score, the greater the death anxiety (Tomás-Sábado et al., 2005). In the present study, the dichotomous version (yes or no) was used because it would be preferred and more effortless for aged adults to respond (Quinn, 2010). There is consensus on their minimal impact on validity and reliability if an item offers between two and five response options (Lopez, 2005). Furthermore, the dichotomous option can perform acceptably with smaller sample sizes, as in the present study (Gómez-Benito et al., 2010).

### Procedure

A research assistant instructed participants on the study’s objectives, how to complete demographic information, and the DAI-R during the first quarter of 2022. Previously, the cognitive state when completing the questionnaire was quantified with the Mini-Mental Examination (Folstein et al., 1975). Only participants with scores in the normative range, without neurocognitive decline, for the Colombian population were included in the study (Rosselli et al., 2000).

### Data analysis

A confirmatory factor analysis (CFA) was performed for the four-dimensional version proposed by Tomás-Sábado et al. (2005). The goodness-of-fit coefficients were observed. The proposed dimensionality for the DAI-R would be accepted if at least three goodness-of-fit indicators were within the recommended parameters (Hair et al., 2006; Hu & Bentler, 1999): chi-square test, with degrees of freedom (df), value p, the relationship between the chi-square/df (normalized chi-square) less than 3.00, root mean square error of approximation (RMSEA) and 90% confidence intervals (CI90%), comparative fit index (CFI), index Tucker-Lewis (TLI) and standardized root mean square residual (SRMR). A probability value greater than 5% is expected in the chi-square, with values less than .06 for RMSEA and SRMR and higher than .89 for CFI and TLI.

Additionally, internal consistency was calculated for each dimension of the DAI-R using Cronbach’s alpha (1951) and McDonald’s omega coefficients (1970). Finally, internal consistency for the full version was only computed for comparison with previous studies. The Jamovi and STATA statistical programs were used.

### Ethical issues

An institutional ethics board reviewed and approved the research project because it was supposed to be a low risk to older adults. The participants signed informed consent following Colombian and international standards for human participation in research, and free-use instruments without registered copyright were used.

## Results

The CFA could not prove the four-dimensional structure of the DAI-R previously reported. The normalized chi-square was observed to be less than 3.00. The other goodness-of-fit indicators showed unacceptable values: Chi-squared was 278.77, df of 113, p < .001, chi-square/df of 2.47, RMSEA of .12 (90%CI .10-.14), CFI of .76, TLI of .72, and SRMR of .09. The principal loadings of the items in each factor are presented in Table 2. It can be seen that item 8 (I would never accept a job in a funeral home) and item 14 (I often think I may have a severe disease) showed loadings lower than .40.

**Table 2.** Dimensionality of the Death Anxiety Inventory-Revised (Loadings).

DEATH ANXIETY INVENTORY-REVISED
| Ítem | Dimension |  |  |  |
| --- | --- | --- | --- | --- |
|  | 1 | 2 | 3 | 4 |
| 1. I get upset when I am in a cemetery. |  | .59 |  |  |
| 2. The certainty of death makes life meaningless. | .70 |  |  |  |
| 3. It annoys me to hear about death. | .51 |  |  |  |
| 4. I find it difficult to accept the idea that it all finishes with death. |  |  | .63 |  |
| 5. I think I would be happier if I ignored the fact that I have to die. | .56 |  |  |  |
| 6. I think I am more afraid of death than most people. | .84 |  |  |  |
| 7. I find it really difficult to accept that I have to die. | .79 |  |  |  |
| 8. I would never accept a job in a funeral home. |  | .14 |  |  |
| 9. The idea that there is nothing after death frightens me. |  |  | .64 |  |
| 10. The idea of death troubles me. |  |  | .74 |  |
| 11. I very often think about the cause of my death. |  |  |  | .71 |
| 12. Coffins make me nervous. |  | .75 |  |  |
| 13. I am worried about what is after death. |  |  | .57 |  |
| 14. I often think I may have a serious disease. |  |  |  | .34 |
| 15. Dying is the worst thing that could happen to me. | .57 |  |  |  |
| 16. The sight of a corpse deeply shocks me. |  | .69 |  |  |
| 17. I frequently think of my own death. |  |  |  | .70 |

Factor 1 (death acceptance) and factor 3 (death finality) showed high correlations between the items: Factor 1 reached a Cronbach’s alpha of 0.81 and a McDonald’s omega of .82, and factor 3 computed a Cronbach’s alpha of .73 and a McDonald’s omega of .74. However, factor 2 (externally generated death anxiety) and factor 4 (thoughts about death) presented low internal consistency values: factor 2 reached a Cronbach’s alpha value of .60, and McDonald’s omega of .65, and factor 4 had a Cronbach’s alpha coefficient of .62. and McDonald’s omega of .67. The full DAI-R showed a Cronbach’s alpha of .87 and a McDonald’s omega of .88.

Due to the failure to corroborate the four-dimensionality of the DAI-R and the poor performance of the internal consistency of factor 2 (externally generated death anxiety) and factor 3 (thoughts about death), different versions with a smaller number of items were tested. The refinement allowed the authors to find a single version of ten items with goodness-of-fit indicators within the recommended parameters. The findings for that version are shown in Annex 1.

## Discussion

The current study was unable to corroborate the four-dimensional structure of the DAI-R among older Colombian adults.

The present sample of older adults discarded the four-dimensional structure proposed early for DAI-R. This observation is divergent from a couple of previous studies. Waite et al. (2022) applied a five-option Likert version of the DAI-R in 2,205 adults between 18 and 83 years old, and Tomás-Sábado et al. (2005) used the Likert version of five response options to 866 professionals and university students between 17 and 51 years old, observing that the data fit the proposed dimensional structure. It is well known that dimensionality can differ depending on the characteristics of the population since response patterns are usually affected by a set of variables (Keszei et al., 2010). Moreover, the available studies have evident cultural differences (Hedrih, 2019). It is necessary to consider that aspects such as the age of the participants (Quinn, 2010) and the number of options for each item can affect the response to a measurement scale; to the extent that the number of options is reduced, it is more likely to observe a deterioration in the validity and reliability of the measurement (Lozano et al., 2008).

Additionally, in the present study, marked differences were found in the internal consistency values of the DAI-R dimensions. Previous CFA underestimated the importance of these values and omitted them from papers (Tomás-Sábado et al., 2005; Waite et al., 2022). Low internal consistency for the factors of a scale usually predicts poor values in the dimensionality goodness-of-fit indicators (Green & Yang, 2015). Furthermore, more orthodox psychometricians maintain that traditional internal consistency coefficients are exclusively appropriate for unidimensional scales (Tavakol & Dennick, 2011).

In the study presented, item 8 (I would never accept a job in a funeral home) was retained in factor 2 (externally generated death anxiety), and item 14 (I often think I may have a severe disease) was located in factor 4 (thoughts about death) showed low loadings and, subsequently, these factors showed low internal consistency, values below .70 (Campo-Arias & Oviedo, 2008; Keszei et al., 2010; Tavakol & Dennick, 2011). The mediocre loadings of items 8 and 14 in the factor analysis accounted for the low internal consistency observed for factors 2 and 4 (Streiner et al., 2015).

Unsurprisingly, the full version of the DAI-R has consistently shown high consistency with some items with low loadings because internal consistency is highly sensitive to the number of items (Campo-Arias & Oviedo, 2008). This fact has been documented repeatedly in studies that have used the DAI-R (Edo-Gual et al., 2015; Limonero et al., 2003; Onu et al., 2021; Tomás-Sábado & Limonero, 2004; Vallés-Fructuoso et al., 2019). The values of Cronbach’s alpha and McDonald’s omega are overestimated when computed with a more oversized item set of 15 (Campo-Arias & Oviedo, 2008; Tavakol & Dennick, 2011).

Death anxiety can partially account for the fears of older adulthood (Benton et al., 2007). Holistic geriatric care should include the evaluation and management of death anxiety (Eliopoulos, 2018). This approach demands instruments with high validity and reliability. For this, it is essential to know the psychometric performance of the scales in different contexts and populations, including older adults. The performance of health measurement instruments can show unexpected results (dimensions with poor internal consistency) for multiple reasons, especially demographic characteristics such as age and cultural background (Campo-Arias & Oviedo, 2008; Keszei et al., 2010; Streiner et al., 2015).

This study had the novelty of exploring the dimensionality of the DAI-R in an exclusive sample of older adults and calculating internal consistency for each dimension as is appropriate for multidimensional psychometric instruments because, in practice, each dimension represents an independent scale, and it is not completely necessary, if desirable, for the dimensions to show a high correlation between them (Davenport et al., 2015; Tavakol & Dennick, 2011). However, this analysis was carried out with a relatively small sample for a CFA. A sample of one hundred participants can be appropriate for factor analyses because only the chi-square test is the most sensitive measure of fit to sample size (Streiner et al., 2015). A small sample can be of high quality if high loadings are observed and a reduced number of dimensions is expected (Singh et al., 2016) or if there are at least five participants for each instrument item (Nunnally & Bernstein, 1994). Moreover, sample sizes can be under 200 hundred participants with a dichotomous option pattern scale (Gómez-Benito et al., 2010). However, more conservative positions maintain that a CFA with 400 or more participants allows more robust conclusive observations due to the more stable sample loadings (Kyriazos, 2018).

It is concluded that the four-dimensional structure of the DAI-R among Colombian older adults needs improvement. It is necessary to corroborate these findings with a large sample of participants. This observation should be taken as preliminary.

## Data Availability

The data supporting this study's findings are available from the corresponding author upon reasonable request.

---

**Annex 1.** Dimensionality of the Death Anxiety Inventory (ten-item version).

| Ítem | Dimension |  |
| --- | --- | --- |
|  | 1 | 2 |
| 2. The certainty of death makes life meaningless. | .70 |  |
| 4. I find it difficult to accept the idea that it all finishes with death. |  | .66 |
| 5. I think I would be happier if I ignored the fact that I have to die. | .56 |  |
| 6. I think I am more afraid of death than most people. | .81 |  |
| 7. I find it really difficult to accept that I have to die. | .84 |  |
| 10. The idea of death troubles me. |  | .77 |
| 11. I very often think about the cause of my death. |  | .70 |
| 13. I am worried about what is after death. | .53 |  |
| 15. Dying is the worst thing that could happen to me. |  | .65 |
| 17. I frequently think of my own death. |  | .70 |
| Factor 1 (Death acceptance): Items 2, 5, 6, 7, and 13. Cronbach's alpha of .81, and McDonald's omega of .82. |  |  |
| Factor 2 (Death finality): Items 4, 10, 11, 15, and 17. Cronbach's alpha of .83 and McDonald's omega of .83. |  |  |
| Chi-squared of 48.37, df of 34, $p = .052$, normalized chi-square of 1.42, RMSEA of .06 (90%CI .01-.10), CFI of .96, TLI of .95, and SRMR of .04. | | |

